# UPTAKE OF REPRODUCTIVE, MATERNAL AND NEWBORN HEALTH IN THE CONTEXT OF COVID-19 PANDEMIC IN KENYA

**DOI:** 10.1101/2024.01.18.24301357

**Authors:** Joyce Jebet, Ruth Muia, Abednego Ongeso, Blasio Omuga, Grace Omoni, Miriam Wagoro

**Author notes:** Corresponding author: Joyce Jebet.

## Abstract

**Background:** The outbreak of COVID 19 in 2019 lead to destabilization of all sectors globally including access to health care. The strain on the health care system as a result of the disease outbreak led to a shift in operations in the health care system. Maternal and neonatal care was affected as women and their families could not freely access health care owing to the restrictions to curb the spread of COVID 19. This led to a risk of a reversal in the gains made in maternal and neonatal health, prompting the need to strengthen community midwifery.

**Aim:** The aim of this study was to strengthen community reproductive, maternal and newborn health in the context of COVID-19 pandemic in Kenya.

**Materials and methods:** This is part of an interventional study that sought to strengthen community maternal and neonatal health services. A baseline survey was conducted to assess the uptake of reproductive, maternal and neonatal care services in one sub-county in Kilifi and Kitui, where five wards in each sub-county were selected. An interviewer administered questionnaire was used to collect data. The sample size for the two Counties was 414.

**Results:** There was a total of 378 respondents mostly comprising a rural population 75.7% (n=286). Outpatient and COVID 19 services (tests and vaccine) were most sought 37.5% (n= 137) and 29.9% (n=109) respectively. Antenatal care services were sought by 26% (n=95) of the respondents, postnatal care 9% (n=33) and skilled birth attendance 8.8% (n=32).

**Conclusion:** The most utilized health facility services were outpatient and COVID 19 services. Antenatal care was also sought, however skilled birth attendance and postnatal care services were least sought.

## Introduction

Maternal and neonatal mortality is still a major public health concern in developing nations. According to the Kenya Demographic Health Survey (1) report, Kenya’s maternal mortality ratio (MMR) of 362 per 100,000 live births and 23 stillbirths per 1,000 was way above the Sustainable Development Goal (SDG) set target of MMR of 70 per 100,000 and 12 stillbirths per 1,000 live births (2).

While the health facility delivery rates had increased from 43% in the year 2008/09 to 62% in 2014 and 75% in 2022 according to the Kenya demographic Health Survey (KDHS) reports (3)(4), the country has challenges in moving towards attaining the Sustainable Development Goal (SDG) 3 which aims at reducing maternal mortality ratio to less than 70 women per 100,000 live births, neonatal mortality rate to less than 12 per 1,000 live births. These challenges are confounded by the COVID -19 pandemic that has affected access to for maternal and neonatal health (MNH) services. According to (5), there were no significant changes on ANC attendance, Facility births, FP uptake and PNC attendance among women, however the study reported an increase in teenage pregnancy. Fresh still births increased with a marked increase in FP uptake among adolescents to include increase in Caesarean Sections among adolescents. These declining indicators have been associated with government-imposed travel restrictions, curfews, loss of many families’ income and fear about COVID infections among clients seeking Reproductive, Maternal and Neonatal Health (RMNH) services in health facilities. These factors poor indicators and service disruptions are likely to adversely affect the gains made in provision of Maternal and Neonatal Health services (MNH) in the country if no strategic interventions are instituted. With most published studies on COVID 19 and RMNH in Kenya being qualitative in nature, data on current trends on RMNH indicators is scanty.

To address MNH needs in the context of COVID 19, the government of Kenya developed guidelines to guide the provision of RMNH services during this period. In addition, the National Guidelines for continuity of Community Health Services was as developed to give policy direction on management of the health services at the Primary Health care level during the pandemic. Development of these Guidelines was the step towards the right direction as it was in tandem with the move towards the achievement of Universal Health Coverage by the year 2030. The Kenya country is committed to achieve the UHC through strategic strengthening of the Primary Health Care system at the community level with particular focus on Kenya Essential Package of Health (KEPH) Levels 1-3 (individuals, Households, Dispensaries and health Centres). Moreover, Universal Health Coverage was one of the presidential Flagship projects reflected on the big 4 agendas of the current government of Kenya to be institutionalized by the year 2022. Nevertheless, the in RMNH guidelines (2020) does not comprehensively address the continuity of the Reproductive, Maternal and Neonatal Health (RMNH) services in the context of COVID19 at the community level. In addition, the continuity of community health services in COVID19 situation guidelines (2020) don’t show an implementation framework of RMNH service provision.

Strengthening the community reproductive, maternal and newborn health services is in line with Kenya health policy framework and SDG 3 and therefore adapting an appropriate model that has proven potential to enhance access to RMNH will guide policy direction on this area. This will provide an Impetus for reviewing/revising the existing RMNH Guidelines and the community Health guideline with a view to strengthening access to Reproductive, Maternal and Newborn Health (RMNH) services at the community level. Therefore, this intervention is timely and is likely to be highly supported if it is successful in the context of strengthening primary Health care towards achievement of Universal health Coverage making country replication feasible and much easier. The goal of the project was improve access to community RMNH services in the context of COVID -19 pandemic in Kenya.

## Materials and Methods

This was a mixed method implementation research that utilized both quantitative study design. It was implemented through a cyclic process of implementing and learning to improve the practice. The implementation sites were Kilifi and Kitui Counties, which were purposively selected owing to their high burden for maternal and neonatal health and availability of a robust community health system. A baseline survey was conducted focusing on selected community health units and the link health facilities to determine the disruptions of health services at the onset and during COVID 19 pandemic.

Therefore, gap analysis baseline survey with a focus to service uptake of Reproductive, Maternal and Neonatal Health (RMNH) Service at the level of County Health Management Teams (CHMT)/ sub-County CHMT, facility service managers and service providers was conducted. Baseline assessment of the facility was conducted using a facility assessment tool, Focus Group Discussions (FGDs) with the service providers and Client exit Interviews was used.

## Results

### Sociodemographic characteristics of the study participants

The study enrolled 378 females aged between 16 and 59 years from Kilifi and Kitui counties (46.8% and 53.2%). Their sociodemographic characteristics are outlined in Table 1. The median (interquartile range) age of the study participants was 27.0 (23.0 - 38.0) years. Those who were aged between 21-30 years were 47.4% while those between the age of 31 and 40 years were 17.2%. The majority were married (65.6%), Christians (92.1%), Self-employment (72.2%) and hailed from rural areas (75.7%). Analysis of the educational background of the study participants revealed that those who had primary and secondary level education were 32.8% and 40.5% respectively.

**Table 1:**
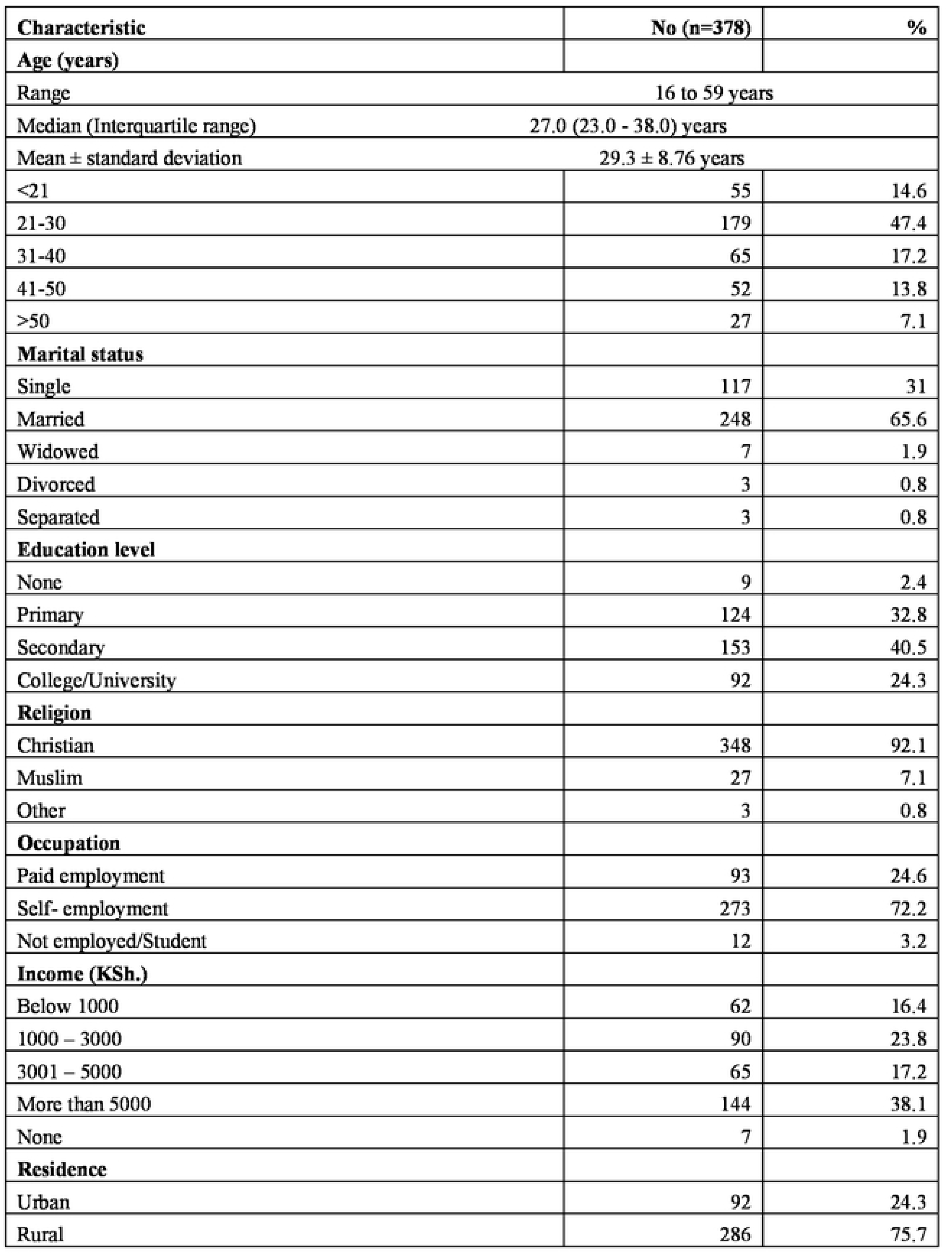
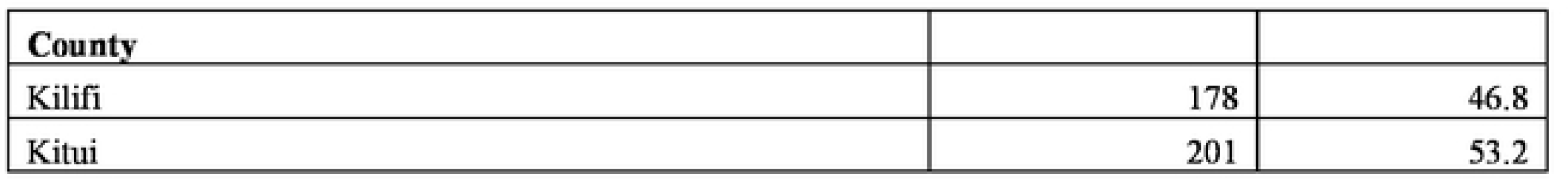
Sociodemographic characteristics of the study participants.

| **Characteristic** | **No (n=378)** | **%** |
| --- | --- | --- |
| **Age (years)** |  |  |
| Range | 16 to 59 years |  |
| Median (Interquartile range) | 27.0 (23.0 - 38.0) years |  |
| Mean ± standard deviation | 29.3 ± 8.76 years | |
| <21 | 55 | 14.6 |
| 21-30 | 179 | 47.4 |
| 31-40 | 65 | 17.2 |
| 41-50 | 52 | 13.8 |
| >50 | 27 | 7.1 |
| **Marital status** |  |  |
| Single | 117 | 31 |
| Married | 248 | 65.6 |
| Widowed | 7 | 1.9 |
| Divorced | 3 | 0.8 |
| Separated | 3 | 0.8 |
| **Education level** |  |  |
| None | 9 | 2.4 |
| Primary | 124 | 32.8 |
| Secondary | 153 | 40.5 |
| College/University | 92 | 24.3 |
| **Religion** |  |  |
| Christian | 348 | 92.1 |
| Muslim | 27 | 7.1 |
| Other | 3 | 0.8 |
| **Occupation** |  |  |
| Paid employment | 93 | 24.6 |
| Self- employment | 273 | 72.2 |
| Not employed/Student | 12 | 3.2 |
| **Income (KSh.)** |  |  |
| Below 1000 | 62 | 16.4 |
| 1000 – 3000 | 90 | 23.8 |
| 3001 – 5000 | 65 | 17.2 |
| More than 5000 | 144 | 38.1 |
| None | 7 | 1.9 |
| **Residence** |  |  |
| Urban | 92 | 24.3 |
| Rural | 286 | 75.7 |

| County |  |  |
| --- | --- | --- |
| Kilifi | 178 | 46.8 |
| Kitui | 201 | 53.2 |

### Uptake of community RMNH services

A total of 221 respondents (58.5%) responded they or their family sought RMNH services during the COVID-19 era. ANC, PNC, CWC, FP and maternity services were sought by 29.5%, 14.5%, 30.7%, 10.8% and 13.3% of the respondents respectively. The service providers for RMNH services as reported by the respondents included nurses and community midwives (89.1% n=337), Community Health Volunteer (4.0% n=15) and Community Health Assistants (0.5%). Other respondents sought services from other providers who included herbalists, relatives and traditional healers (1.9% n=7), whereas others did self-medication (4.5% n=17)

A total of 41 respondents (10.8%) changed their provider of RMNH services from during the COVID-19 period. The reasons provided for the change included fear of getting COVID-19 infection 43.9%), health facility being designated as an isolation centre (14.6%), transport constraints (14.6%) and lack of pertinent information from the CHV (7.3%). Other reasons included unavailability of the healthcare workers (4.9%), long distance to the health centre (4.9%), and Financial constraints (4.9%).

Most of the study participants reported public facilities as their current health facility for seeking RMNH services and also preferred type of health facility for seeking RMNH services (95.2% and 92.3% respectively). Those who were seeking RMNH services in private health facilities at the time of conducting the study were 4.2%. Additionally, 7.1% reported that they preferred seeking RMNH services in private health facilities.

**Figure 1:**
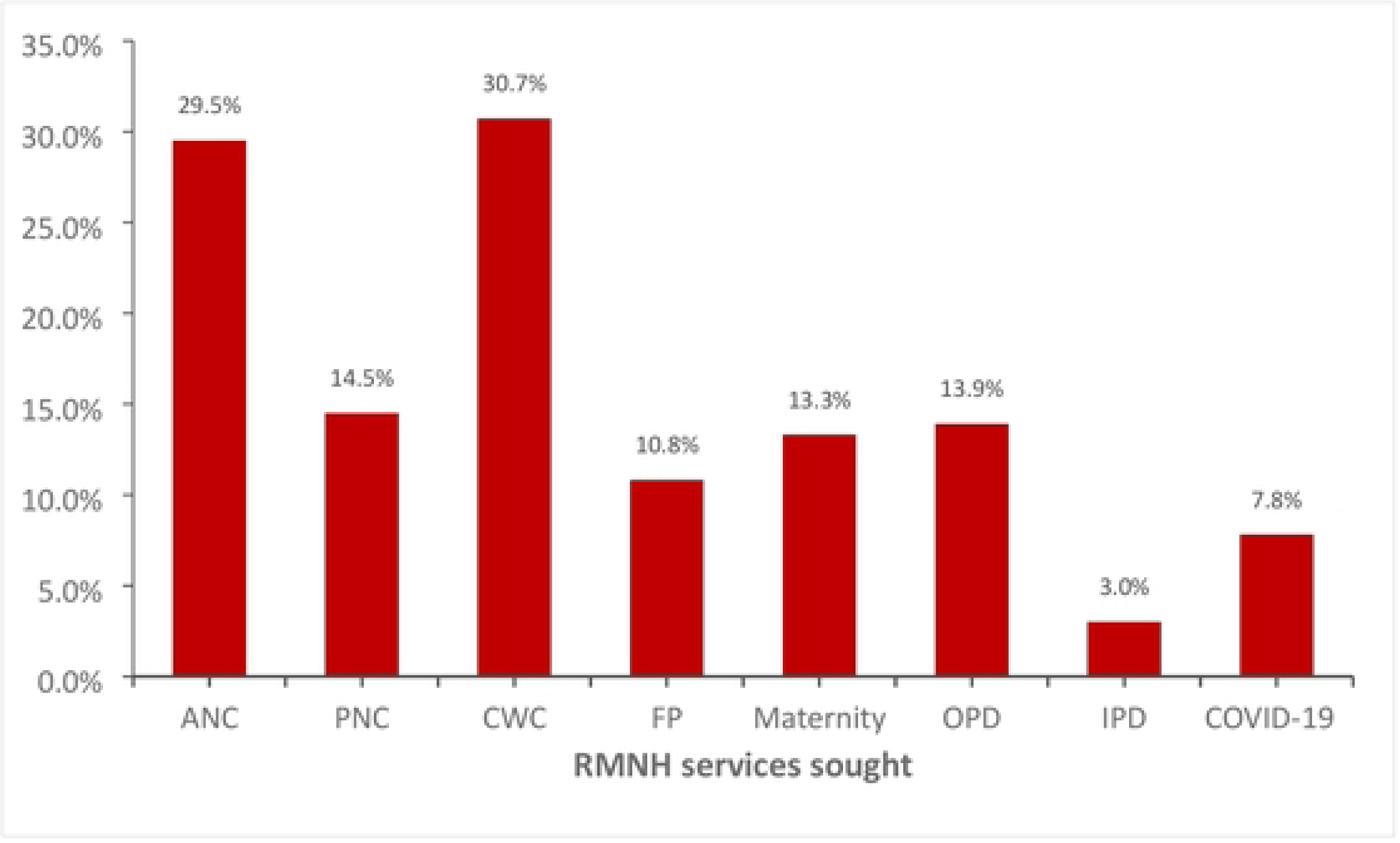
Services sought during COVID-19 era.

### Uptake of community RMNH services

The health care rated the uptake of RMNH services before the onset of COVID 19 as higher (57.1%) or lower (42.9%). However during the peak of COVID 19 in 2020, majority (90%) rated the services as disrupted. Most of them (63.3%) agreed that the services uptake in 2021 (post-COVID 19) were higher.

**Table 2:** Rating the service uptake for RMNH pre- and post-COVID 19.

| Question | Category | Count | Column N % |
| --- | --- | --- | --- |
| How would you rate the service uptake for RMNH before the onset of COVID 19? | Higher | 16 | 57.1% |
|  | The same as now | 0 | 0.0% |
|  | Lower | 12 | 42.9% |
| How would you rate the service uptake for RMNH in the peak period of COVID in 2020? | Service uptake not affected | 2 | 6.7% |
|  | Services disrupted | 27 | 90.0% |
|  | Higher service uptake | 1 | 3.3% |
| Q19 How would you rate the service uptake for RMNH currently in 2021 compared to 2020? | Higher | 19 | 63.3% |
|  | The same as now | 3 | 10.0% |
|  | Lower | 8 | 26.7% |

### Reasons for not seeking RMNH services during the COVID-19 era

Most of the respondents (61.3%) reported that they were not sick/did not require RMNH services during the period under enquiry. Those who failed to seek RMNH services for fear of contracting COVID-19 infection in the health facilities were 3.9% while 1.2% reported that COVID-19 restrictions prevented them from seeking the services.

**Table 3:** Reasons for not seeking RMNH services.

| **Reason** | **Number (n=155)** | **%** |
| --- | --- | --- |
| Not sick | 95 | 61.3 |
| Fear of contracting COVID-19 in the health facilities | 6 | 3.9 |
| Had information that no drugs were available | 3 | 1.9 |
| Visiting the herbalist | 2 | 1.3 |
| Not aware of the services | 2 | 1.3 |
| Stigma and fear/distance and fear/Presence of herbal doctors and midwives | 2 | 1.3 |
| The facility is far | 2 | 1.3 |
| Advanced age | 2 | 1.3 |
| COVID-19 protocols prevented | 2 | 1.3 |
| Don't Know | 2 | 1.3 |

### Barriers that hindered utilization of RMNH services in the community

The reported barriers included fear of acquiring COVID-19 infection (38.6%), lack of knowledge (16.7%), religion (8.7%), financial difficulties (5.6%) and unfavourable treatment from nurses (4.8%).

### Opportunities for improving RMNH services in the community

These included increasing the number of health facilities (33.6%), bringing services to the people (18.8%), providing access to contraceptives and safe delivery (10.4%) and providing access to contraceptives and safe delivery (5.8%) (Table 4)

**Table 4:** Barriers that prevent utilization of RMNH services in the community.

| **Barrier** | **No (n=378)** | **%** |
| --- | --- | --- |
| Fear of acquiring COVID-19 infection | 146 | 38.6 |
| Lack of knowledge and understanding of the importance of services/poor health education | 63 | 16.7 |
| False information, misconceptions, myths, misbeliefs etc. | 33 | 8.7 |
| Religion | 21 | 5.6 |
| Financial difficulties | 18 | 4.8 |
| Bad treatment from nurses/ health workers | 16 | 4.2 |
| Long-distance to health facilities | 15 | 4.0 |
| Traditional beliefs | 6 | 1.6 |
| Side effects of contraceptives | 4 | 1.1 |
| Transport challenges | 4 | 1.1 |
| Others | 52 | 13.8 |

Among the suggestions to improve uptake of RMNH services were health education/promotion (34.4%), increment of health facilities (15.3%), recruitment of more health care workers (11.6%), raising awareness in the community (11.1%), and empowerment of women (8.2%) (Table 5).

**Table 5:**
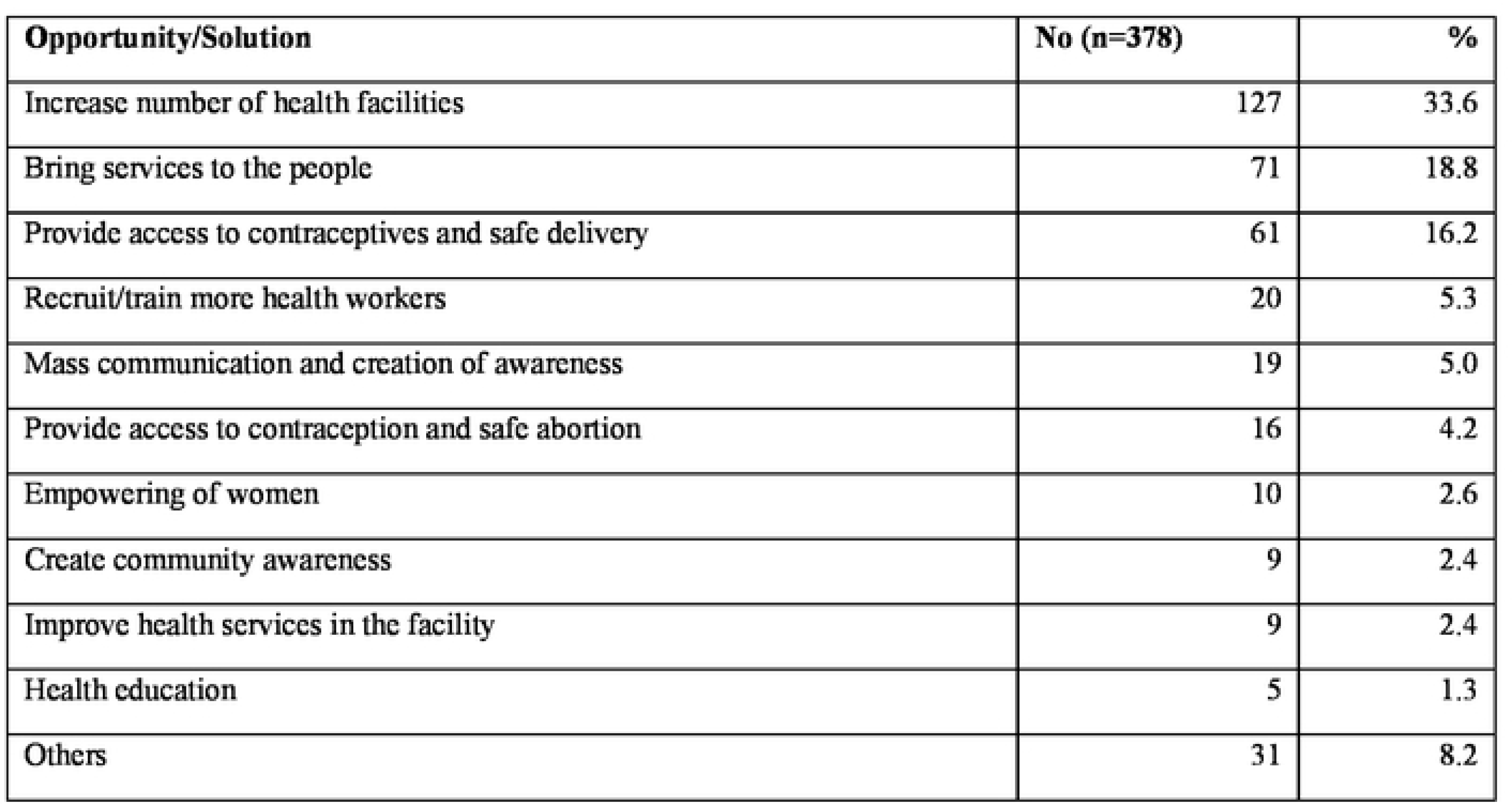
Opportunities for improving RMNH services in the community.

## Discussion

The respondents were aged between 16 to 59 years with a median of 27 years. This indicates that majority were a younger population. This could be explained by the Kenya’s expanding pyramid population where there are younger people at the bottom. This age category is also the group which is economically productive and also in reproductive matters. Age affects economic productivity in that with increasing age, there is reduced economic productivity (6).

The study population was a rural community with low socio-economic status evidenced by their monthly average income of less than 5,000 Kenyan shillings per month. In addition, majority of them had primary or secondary level of education as their higher achievement. There is a strong relationship between level of education and income where, a higher level of education is linked to a higher income (7,8).

The most sought health facility services were outpatient department services, child welfare services and COVID 19 services which included tests and vaccines. However, maternity services for skilled birth attendance and postnatal care (PNC) were least sought though antenatal care (ANC) was highly sought. This could indicate that there was a likelihood of homebirths during the time and also ignorance on postnatal care services. Although studies (9)(10) have established that women who attend ANC are more likely to have a hospital birth and subsequently seek PNC services, this study established the contrary. This could be attributed to the fear of contracting COVID 19 at the time due to rumours and misconceptions that had not been dispelled. Other reasons for not seeking care in health facilities included distance, stigma and availability of herbalists.

Access to RMNH was affected by many barriers during the COVID 19 pandemic. In the current study, barriers to accessing RMNH ranged from fear of acquiring COVID 19, lack of knowledge, false information, misconceptions and myths about COVID 19 and religion. Financial and transport challenges was also a barrier. Negative attitude of health care workers also contributed to clients not seeking services. Similar findings were established in other studies (11)(12). Other than lock down and restriction of movement, distance to the health facilities also contributed as a barrier to accessing reproductive health services. This was not only unique to Kenya but other countries as well (11,13–15).

The barriers during the COVID 19 pandemic opens a discussion on what could be done during the pandemic. Some of the opportunities as suggested by the respondents include creating awareness to the community, women empowerment, mass media communication and bringing services to the people. In addition, the findings of this study posits the need to utilize technology to bridge the gap of access to services. To address the gap of distance to the health facilities, there is need to have equity in distribution of the health facilities. This calls for concerted efforts by governments to address this and prevent similar challenges in case of other pandemic situations (14).

## Conclusions

Uptake of reproductive, maternal and neonatal health services were affected during the COVID 19 pandemic. Several barriers including fear of contracting COVID 19, stigma, perception on lack of services and negative attitude of health care providers contributed to low uptake of services.

## Data Availability

All data produced in the present work are contained in the manuscript

## Acknowledgments

We acknowledge the United Nations People’s Fund (UNFPA) for their financial support towards this project. We also acknowledge AMREF for administration of the funds. In addition, we acknowledge Kilifi and Kitui County management for actively supporting the project.

## Other elements

Tables and figures are uploaded separately.

